# Infection prevention and control preparedness among public hospitals and COVID-19 temporary treatment and monitoring facilities in the Philippines

**DOI:** 10.1101/2022.05.11.22274966

**Authors:** Vergil de Claro, Noemi Bautista, Ma. Rosario Torralba, Vina Vanessa Castro, Miguel Angelo Lucero, Lady Jedfeliz Molleno, Laurentiu Stan

## Abstract

**Objectives:** An effective response to COVID-19 necessitates rigid compliance of health facilities to infection prevention and control (IPC) protocols to protect HCWs, prevent onward transmission, and mitigate the impact of the outbreak on the health care system. The study aims to assess the compliance of public hospitals and temporary treatment and monitoring facilities (TTMFs) to IPC standards for COVID-19.

**Methods:** A baseline assessment survey was conducted between July 20 to August 18, 2020, from selected facilities in 38 local government units (LGU) across the country utilising a 35-point questionnaire corresponding to a set of standards issued by the Philippine Department of Health.

**Results:** The study shows that public hospitals reported sufficient IPC preparedness and compliance compared to TTMFs in the domains of engineering and administrative controls. Both types of facilities reported weak compliance in the use of color-coded bags and in having a central storage for infectious waste. In addition, among TTMFs adherence to proper labelling of waste bins, presence of instructional materials for waste disposal, directional signages for movements of HCWs and patients, availability of an IPC policy, and advocacy materials on proper handwashing, respiratory etiquette, and physical distancing, and PPE use were also low.

**Conclusion:** The COVID-19 pandemic has shown the importance of IPC preparedness among health care facilities to effectively prevent disease transmission and mitigate its impact on the health care system. The findings suggests that periodic monitoring and augmentation of resources are needed to immediately address the compliance gaps. However, systemic improvements and long-term investments are required to sustain IPC practices over time.

**What is already known?:** Infection prevention and control measures are effective at protecting patients and health care workers from facility-acquired infections and averting onward transmission of the disease.

**What are the new findings?:** Findings from this study highlight the critical gaps in infection prevention and control preparedness among established healthcare settings like hospitals and in repurposed spaces such as temporary facilities for COVID-19 isolation that were primarily set up to manage the surge in cases.

**What do the new findings imply?:** It focuses attention on the periodic monitoring of health facilities’ compliance to standard infection prevention and control practices especially during outbreak situations as a basis for identifying immediate resource and technical requirements, and for planning the needed investments in the long term.

## INTRODUCTION

Coronavirus disease 2019 (COVID-19) which first emerged in Wuhan, China was confirmed to be transmitted from person to person(1). The first reported case of COVID-19 in the Philippines was on January 22, 2020(2) which has now infected 3.06 million Filipinos, 6.8% of which are still active cases(3). There is a greater risk of exposure to COVID-19 among health care workers (HCW) who are at the forefront in responding to the current pandemic(4). This has implications on the capacity of health care facilities to attend to the needs of patients, further exacerbating the inadequacy of the health human resources of an already overburdened health care system.

Globally, 3.9% of COVID patients are reported to be HCWs(5). Early local studies have shown that the risk of exposure to the disease is high among HCWs(6). A single-centre retrospective analysis based on admitted COVID-19 individuals revealed that 26.6% of these patients are HCWs (7). As of December 30, 2021, the Department of Health (DOH) recorded 28,744 HCWs who were positive for COVID-19. Of these, 99.3% (28,539) have recovered, 0.4% (115) have died and 3.2% (372) are still in critical condition(3).

Current evidence demonstrates the effectiveness of infection prevention and control (IPC) measures including the use of personnel protective equipment (PPE), disinfection practices, engineering, and environmental controls. National IPC guidelines existed prior to COVID-19 and are part of the regulatory requirements for health care facilities being granted a license to operate. These standards(8-10) were updated by DOH in line with the recommendations from the World Health Organization (WHO) for COVID-19(11). The level of compliance, however, with the updated IPC measures remains unknown. An assessment of how these standards have been adopted and complied with provides information for addressing any gaps and in further strengthening IPC practices in health facilities with the objectives of reducing disease transmission and mitigating the impact of the ongoing and future outbreaks.

## METHODS

We assessed the compliance to COVID-19 IPC standards of public hospitals and COVID-19 temporary treatment and monitoring facilities (TTMFs) based on the guidelines issued by the DOH. The assessment was conducted between July 20 to August 18, 2020, from selected facilities in 38 local government units. Public hospitals and TTMFs were selected through convenience sampling across the three main island groups of the country – Luzon, Visayas, and Mindanao.

The primary objective of the assessment was to gain rapid insight into the preparedness of healthcare facilities in implementing IPC standards and identifying immediate areas of support during the pandemic. The survey utilises a 35-point questionnaire corresponding to a set of standards for the following domains: 1) engineering controls, 2) environmental controls, 3) waste management, and 4) administrative controls. The tool was administered by trained field data collectors either virtually through phone interviews using Zoom and/or Facebook Messenger, or face-to-face meetings following strict adherence to public health safety protocols. Responses were directly encoded into pre-designed Google Forms together with photos or video clips as means of verification (MOVs). Descriptive statistics were used to analyse the survey data. Differences between the island groups were tested using Pearson’s χ^2^ test for categorical variables.

### Public Involvement

Respondents were not involved in the design and management of the study and their participation was limited only to providing information related to policies, procedures, and practices within their respective institutional settings. However, local decision-makers were informed of the baseline results so that necessary support can be mobilised from the concerned local government units and their partners.

## RESULTS

A total of 222 responses were received comprising of 83 (37.4%) public hospitals and 139 (62.6%) TTMFs. Combined responses were collected from across the main island groups of Luzon 89 (40.1%), Visayas 36 (16.2%), and Mindanao 97 (43.7%) which translates to a nationwide representation for both types of facilities in this baseline assessment. The demographic characteristics of the health facilities and the respondents are shown in Table 1.

**Table 1.** Demographic characteristics of health facilities and respondents.

| Characteristics | No. of Respondents<br>n (%) | Comparison between island groups |  |  |
| --- | --- | --- | --- | --- |
|  |  | Luzon<br>n (%) | Visayas<br>n (%) | Mindanao<br>n (%) |
| *Public hospitals* | 83 (100.0) | 39 (46.99) | 15 (18.07) | 29 (34.94) |
| Respondents by profession |  |  |  |  |
| Chief of Hospital | 9 (10.8) | 2 (5.1) | 3 (20.0) | 4 (13.8) |
| IPC Physician | 3 (3.6) | 2 (5.1) | 1 (6.7) | - |
| IPC Nurse | 53 (63.9) | 26 (66.7) | 6 (40.0) | 23 (79.3) |
| IPC Officer | 7 (8.4) | 4 (10.3) | 3 (20.0) | - |
| Other professionals | 11 (13.3) | 5 (12.8) | 2 (13.3) | 2 (6.0) |
| DOH certified | 85 (100.0) | 39 (100.0) | 15 (100.0) | 29 (100.00) |
| PhilHealth accredited | 78 (93.9) | 39 (100.0) | 14 (93.3) | 25 (86.2) |

| *TTMFs* | 139 (100.0) | 50 (35.97) | 21 (15.11) | 68 (48.92) |
| --- | --- | --- | --- | --- |
| Respondents by profession |  |  |  |  |
| Facility Manager | 32 (23.02) | 6 (18.75) | 4 (12.50) | 22 (68.75) |
| IPC Physician | 25 (17.99) | 9 (36.0) | 5 (20.0) | 11 (44.0) |
| IPC Nurse | 49 (35.25) | 18 (36.73) | 10 (20.41) | 21 (42.86) |
| Other professionals | 33 (23.74) | 17 (51.52) | 2 (6.06) | 14 (42.42) |
| DOH certified | 76 (54.29) | 24 (32.0) | 13 (17.33) | 38 (50.67) |
| PhilHealth accredited | 27 (19.29) | 7 (25.93) | 13 (48.15) | 7 (25.93) |

IPC, infection prevention and control; DOH, Department of Health; TTMF, temporary treatment and monitoring facility

The results presented in Table 2 and Table 3 show that public hospitals are highly compliant with standards in the domains of engineering, and administrative controls. Conversely, for the waste management domain, only 104 of the 222 surveyed facilities have reported the use of color-coded bags [46.8%; confidence interval (CI): 45.8–47.8] showing weak compliance by hospitals [69.9%; 95% CI: 68.7–71.1] and TTMFs [33.1%; 95% CI: 32.4–33.8] alike. Similarly, for both types of facilities, 33% reported not having a central storage for infectious wastes with only 58 of 83 hospitals [69.9%; 95% CI: 68.7–71.1] and 97 of 139 TTMFs [69.8%; 95% CI: 67.3–72.3] are compliant.

**Table 2.** A geographical comparison of public hospitals’ compliance to IPC protocols, survey results from 83 responses in the Philippines, August 4-18, 2020.

| Standards | No. of Respondents<br>n (%) | Comparison between island groups |  |  |  |
| --- | --- | --- | --- | --- | --- |
|  |  | Luzon<br>n (%) | Visayas<br>n (%) | Mindanao<br>n (%) | p-value |
| Engineering Controls |  |  |  |  |  |
| Triage area has hand hygiene area | 81 (97.6) | 39 (100.0) | 15 (100.0) | 26 (89.7) | 0.005 |
| Triage area have directional signages | 66 (79.5) | 35 (89.7) | 14 (93.3) | 17 (58.6) | 0.003 |
| HCWs wearing proper PPE | 80 (96.4) | 39 (100.0) | 15 (100.0) | 26 (89.7) | 0.005 |
| Cleaners wearing proper PPE | 79 (95.2) | 37 (94.9) | 15 (100.0) | 27 (93.1) | 0.010 |
| Emergency room isolation has contaminated zone | 76 (91.6) | 37 (94.9) | 14 (93.3) | 25 (86.2) | 0.005 |
| Emergency room isolation has a buffer zone | 76 (91.6) | 37 (94.9) | 15 (100.0) | 24 (82.8) | 0.008 |
| Emergency room isolation has sterile zone | 74 (89.2) | 37 (94.9) | 13 (86.7) | 24 (82.8) | 0.003 |
| COVID-19 isolation ward has contaminated zone | 71 (85.5) | 36 (92.3) | 14 (93.3) | 21 (72.4) | 0.005 |
| COVID-19 isolation ward has buffer zone | 71 (85.5) | 36 (92.3) | 14 (93.3) | 21 (72.4) | 0.005 |
| COVID-19 isolation ward has sterile zone | 70 (84.3) | 36 (92.3) | 13 (86.7) | 21 (72.4) | 0.003 |
| Environmental Controls |  |  |  |  |  |
| Cleaning and disinfection of surface areas once a day | 82 (98.8) | 38 (97.4) | 15 (100.0) | 29 (100.0) | 0.007 |
| Cleaning and disinfection upon discharge of patient | 82 (98.8) | 39 (100.0) | 15 (100.0) | 28 (96.6) | 0.005 |
| Use of 70% ethyl alcohol or 0.1% sodium hypochlorite to disinfect surfaces | 81 (97.6) | 39 (100.0) | 15 (100.0) | 27 (93.1) | 0.005 |
| Use of 0.5% sodium hypochlorite to clean bodily fluids | 78 (93.9) | 38 (97.4) | 15 (100.0) | 25 (86.2) | 0.006 |
| Compliance to ≥30 minutes standard waiting time for disinfection | 82 (98.8) | 39 (100.0) | 15 (100.0) | 28 (96.6) | 0.005 |
| Waste Management |  |  |  |  |  |
| Appropriate labelling of waste bins | 79 (95.2) | 39 (100.0) | 15 (100.0) | 27 (93.1) | 0.005 |
| Use of color-coded bags | 58 (69.9) | 27 (69.2) | 15 (100.0) | 16 (55.2) | 0.101 |
| Presence of posters/printed instructions for disposal | 69 (83.1) | 36 (92.3) | 14 (93.3) | 19 (65.5) | 0.003 |
| Designated temporary collection point for infectious wastes | 76 (91.6) | 37 (94.9) | 13 (86.7) | 28 (96.6) | 0.004 |
| Temporary collection point is covered/sealed | 74 (89.2) | 37 (94.9) | 13 (86.7) | 26 (89.7) | 0.003 |
| Temporary collection point is far from public access | 79 (95.2) | 39 (100.0) | 15 (100.0) | 27 (93.1) | 0.002 |
| Central storage for infectious waste | 58 (69.9) | 27 (69.2) | 15 (100.0) | 16 (55.2) | 0.005 |
| Central storage is proximate to exit gate/<br>garbage pick-up | 69 (83.1) | 36 (92.3) | 14 (93.3) | 19 (65.5) | 0.004 |
| *Administrative Controls* |  |  |  |  |  |
| Written policy on IPC | 74 (89.2) | 37 (94.9) | 13 (86.7) | 26 (89.7) | 0.005 |
| Dedicated IPC Officer | 75 (90.4) | 37 (94.9) | 12 (80.0) | 28 (96.6) | 0.007 |
| Trained HCWs on IPC | 81 (97.6) | 39 (100.0) | 15 (100.0) | 29 (100.0) | 0.007 |
| Familiarity with the steps to proper handwashing | 78 (94.0) | 38 (97.4) | 14 (93.3) | 28 (96.6) | 0.007 |
| Familiarity with the steps to proper donning of PPEs | 81 (97.6) | 38 (97.4) | 15 (100.0) | 28 (96.6) | 0.007 |
| Familiarity with the steps to proper doffing of PPEs | 81 (97.6) | 38 (97.4) | 15 (100.0) | 28 (96.6) | 0.007 |
| Promotional/educational materials on proper<br>handwashing | 74 (89.2) | 39 (100.0) | 15 (100.0) | 20 (86.9) | 0.002 |
| Promotional/educational materials on<br>respiratory etiquette and physical distancing | 72 (86.7) | 32 (82.1) | 15 (100.0) | 25 (86.2) | 0.048 |
| Promotional/educational materials on use of<br>PPEs per zone | 66 (79.5) | 33 (84.6) | 14 (93.3) | 19 (65.5) | 0.012 |

IPC, infection prevention and control; HCW, healthcare worker; PPE, personnel protective equipment; COVID-19, coronavirus disease 2019

**Table 3.** A geographical comparison of TTMFs’ compliance to IPC protocols, survey results from 139 responses in the Philippines, July 20 to August 15, 2020.

| Standards | No. of Respondents<br>n (%) | Comparison between island groups |  |  |  |
| --- | --- | --- | --- | --- | --- |
|  |  | Luzon<br>n (%) | Visayas<br>n (%) | Mindanao<br>n (%) | p-value |
| *Engineering Controls* |  |  |  |  |  |
| Entrance and exit of HCWs is connected to clean/sterile area | 116 (83.5) | 47 (94.0) | 20 (95.2) | 49 (72.1) | <0.001 |
| Entrance and exit of HCWs have directional signages | 68 (48.9) | 29 (58.0) | 14 (66.7) | 25 (36.8) | 0.069 |
| Entrance and exit of patients are connected to contaminated area | 115 (82.7) | 45 (90.0) | 19 (90.5) | 51 (75.0) | <0.001 |
| Entrance and exit of patients have directional signages | 68 (48.9) | 30 (60.0) | 14 (66.7) | 24 (35.3) | 0.056 |
| Use of the following PPE by HCWs: |  |  |  |  |  |
| Face mask | 128 (92.1) | 48 (96.0) | 20 (95.2) | 60 (88.2) | <0.001 |
| Eye protection | 134 (96.4) | 50 (100.0) | 20 (95.2) | 64 (94.1) | <0.001 |
| Gloves | 133 (95.7) | 50 (100.0) | 20 (95.2) | 63 (92.6) | <0.001 |
| Gown | 127 (91.4) | 49 (98.0) | 18 (85.7) | 60 (88.2) | <0.001 |
| Has a designated contaminated zone | 129 (92.8) | 48 (96.0) | 20 (95.2) | 61 (89.7) | <0.001 |
| Has a designated buffer zone | 127 (91.4) | 49 (98.0) | 19 (90.5) | 59 (86.8) | <0.001 |
| Has a designated sterile zone | 124 (89.2) | 47 (94.0) | 19 (90.5) | 58 (85.3) | <0.001 |
| *Environmental Controls* |  |  |  |  |  |
| Cleaning and disinfection of surface areas daily | 134 (96.4) | 49 (98.0) | 19 (90.5) | 66 (97.1) | <0.001 |
| Cleaning and disinfection upon discharge of patient | 139 (100.0) | 50 (100.0) | 21 (100.0) | 68 (100.0) | <0.001 |
| Use of 70% ethyl alcohol or 0.1% sodium hypochlorite to disinfect surfaces | 135 (97.1) | 50 (100.0) | 20 (95.2) | 65 (95.6) | <0.001 |
| Use of 0.5% sodium hypochlorite to clean bodily fluids | 122 (87.8) | 47 (94.0) | 21 (100.0) | 54 (79.4) | <0.001 |
| Compliance to ≥30 minutes standard waiting time for disinfection | 130 (93.5) | 48 (96.0) | 21 (100.0) | 61 (89.7) | <0.001 |
| *Waste Management* |  |  |  |  |  |
| Appropriate labelling of waste bins | 87 (62.2) | 39 (78.0) | 16 (76.2) | 32 (47.1) | 0.008 |
| Use of color-coded bags | 46 (33.1) | 20 (40.0) | 12 (57.1) | 14 (20.6) | 0.323 |
| Presence of posters/printed instructions for disposal | 47 (33.8) | 27 (54.0) | 7 (33.3) | 13 (19.1) | <0.001 |
| Designated temporary collection point for infectious wastes | 114 (82.0) | 46 (92.0) | 18 (85.7) | 50 (73.5) | <0.001 |
| Temporary collection point is covered/sealed | 98 (70.5) | 39 (78.0) | 16 (76.2) | 43 (63.2) | 0.002 |
| Temporary collection point is far from public access | 102 (73.4) | 41 (82.0) | 17 (81.0) | 44 (64.7) | 0.002 |
| Central storage for infectious waste | 97 (69.8) | 42 (84.0) | 15 (71.4) | 40 (58.8) | <0.001 |
| Central storage is proximate to exit gate/garbage pick-up | 102 (73.4) | 42 (84.0) | 17 (81.0) | 43 (63.2) | 0.002 |
| *Administrative Controls* |  |  |  |  |  |
| Written policy on IPC | 82 (58.9) | 25 (50.0) | 10 (47.6) | 47 (69.1) | <0.001 |
| Dedicated IPC Officer | 114 (82.0) | 48 (96.0) | 19 (90.5) | 47 (69.1) | <0.001 |
| Trained HCWs on IPC | 114 (82.0) | 42 (84.0) | 17 (80.9) | 55 (80.9) | <0.001 |
| Familiarity with the steps to proper handwashing | 133 (95.7) | 50 (100.0) | 21 (100.0) | 62 (91.2) | <0.001 |
| Familiarity with the steps to proper donning of PPEs | 132 (94.9) | 48 (96.0) | 21 (100.0) | 63 (92.6) | <0.001 |
| Familiarity with the steps to proper doffing of PPEs | 133 (95.7) | 49 (98.0) | 21 (100.0) | 63 (92.6) | <0.001 |
| Promotional/educational materials on proper handwashing | 78 (56.1) | 35 (70.0) | 18 (85.7) | 25 (36.8) | 0.060 |
| Promotional/educational materials on respiratory etiquette and physical distancing | 58 (41.7) | 27 (54.0) | 14 (66.7) | 17 (25.0) | 0.091 |
| Promotional/educational materials on use of PPEs per zone | 62 (44.6) | 26 (52.0) | 13 (61.9) | 23 (33.8) | 0.106 |

TTMF, temporary treatment and monitoring facility; IPC, infection prevention and control; HCW, healthcare worker; PPE, personnel protective equipment; COVID-19, coronavirus disease 2019

Most TTMFs were also reported not observing standards for proper labelling of waste bins and posting of instructional materials for proper waste disposal within the facility. In the domain of engineering controls, only 68 TTMFs were shown to have directional signages to guide the movement of HCWs and patients [48.9%; 95% CI: 47.6–50.2]. It fell short as well on some sub-set indicators under the administrative domain, specifically on the availability of an IPC policy [58.9%; 95% CI: 55.8–62.0] and advocacy materials for proper handwashing [56.1%; 95% CI: 54.7–57.5], respiratory etiquette and physical distancing [41.7%; 95% CI: 40.6–42.8], and the use of appropriate PPEs per zones [44.6%; 95% CI: 43.5–45.7]. Overall, the baseline assessment reveals that public hospitals reported better IPC preparedness and compliance with standards compared to TTMFs.

## DISCUSSION

Infection prevention and control strategies and measures are essential to prevent COVID-19 transmission in health facilities. Optimal compliance to standards for engineering and environmental control that supports transmission-based precautions such as screening and triage, administrative controls including an IPC policy in place or a dedicated and trained IPC personnel, and proper management of infectious wastes is associated with decreased risk for the spread of the disease(11) and reduced impact on the health system(12). This baseline assessment offers a rapid insight into the IPC preparedness of health facilities during the COVID-19 pandemic and the adaptations necessary to sustain these practices against future outbreaks.

There are evident variations observed between facilities, particularly for engineering and environmental controls with public hospitals reporting a higher degree of compliance over TTMFs. This was expected since available public spaces such as auditoriums, gymnasiums, classrooms, vacant hotels, courts, and even open fields with tents were the ones converted into TTMFs to enhance the surge capacity of existing health facilities(13). Many of these existing spaces must be partitioned into isolation units and fitted with equipment to be functional. Operationally, setting-up engineering controls entail higher initial cost and time compared to procuring PPEs, disinfectants, and health promotional materials as part of regular IPC activities. Indeed, findings from the study have shown that in the early months of the pandemic about half of these facilities were operating not having been certified by the DOH and assessed to have fully complied with IPC and health care waste management protocols among other operational requirements(14) indicating a clear gap in this area.

Another noteworthy observation is the extremely low accreditation rate of TTMFs with PhilHealth, the national health insurance agency. Financing is an essential element for sustaining adherence to IPC measures in health facilities. In situations where the fiscal capacity of health facilities is strained but resources such as availability of health personnel, allocation of PPEs, and utilisation of equipment are continuously demanded and are stretched to the limits, can at some point compromise the practice of IPC. This highlights the need to effectively set a baseline for the resource and infrastructure requirements of temporary facilities on IPC even in non-outbreak settings so that the health system will have a better cushion when infections of COVID-19 like-proportion will happen in the future(15).

Furthermore, a domain of concern is the insufficient disposal capacity of medical waste. Proper segregation, storage, collection, and transport of waste materials in health care facilities catering to infected or suspected patients is critical to ensuring the safety of waste workers and reducing the chance of COVID-19 spreading in the community(16-17). The amount and type of medical waste people in health care settings are exposed to underscore the importance of strictly observing other IPC domains and measures and regularly assessing for their compliance such as the availability of well-defined IPC policies(18), designation of zones(19), appropriate procedure and knowledge in the use of PPEs(20), and well-articulated directional movements of patients and waste materials. We echo the importance of empowering not only health care workers(21) but also engaging the community(22) in practicing IPC guidelines by strengthening individual, organisational, and community-level facilitators and addressing barriers that potentially discourage its sustained implementation.

## CONCLUSION

The COVID-19 pandemic has shown the importance of IPC preparedness among different types of health facilities to effectively prevent disease transmission and mitigate its impact on the health care system. It is particularly important for the local government units to continuously monitor, assist health facilities in meeting the standards, and sustain their compliance over time. The study identifies several compliance gaps that can be immediately responded to through augmentation of resources such as the provision of waste bags, PPEs, signages, and information materials, as well as staff training and technical support in developing institutional IPC policies. Moreover, it also demonstrates the need for systemic improvements and long-term investment in IPC as an essential component in building health systems that are resilient and better prepared for future outbreaks.

## Data Availability

All data produced in the study are available upon reasonable request to the authors.

## Acknowledgments

We are grateful to the contribution of USAID ReachHealth’s provincial and city technical officers who conducted the data collection for this study.

## Contributors

VDC, NB, MRT, and LS proposed and designed the study. NB, VDC and MRT developed the survey instruments. VDC designed the online forms for the encoding of the survey results. VVC, MAL, and LJM oversaw the field data collection and data encoding. VDC and NB prepared the first draft with all authors reviewing and providing input. All authors reviewed and approved the final version of the manuscript.

## Ethics approval

Ethics approval was not required as only information pertaining to institutional policies and processes were collected, with informed consent taken from respondents.

## Data sharing statement

No additional data are available.

